# Essential Workers’ COVID-19 Vaccine Hesitancy, Misinformation and Informational Needs in the Republic of North Macedonia

**DOI:** 10.1101/2021.12.22.21268045

**Authors:** S. Fucaloro, V. Yacoubian, R. Piltch-Loeb, N. Harriman, T. Burmaz, M. Hadji-Janev, E. Savoia

**Affiliations:** Emergency Preparedness Research Evaluation & Practice Program, Harvard T.H. Chan School of Public Health, 90 Smith Street, Boston, 02115 MA; Department of Biostatistics, Harvard T.H. Chan School of Public Health, 99 Huntington Avenue, Boston, 02115 MA; University of Goce-Delcev, Stip, Krste Misirkov 10-A, Shtip 2000, North Macedonia

## Abstract

**Introduction:** The COVID-19 pandemic is a global health crisis that as of December 2021 has resulted in the death of over 5.2 million people. Despite the unprecedented development and distribution of vaccines, hesitancy to take the vaccine remains a wide-spread public health challenge, especially in Eastern European countries. In this study we focus on a sample of essential workers living in the Republic of North Macedonia to: 1) Describe rates of vaccine acceptance, risk perception and sources of COVID-19 information, 2) Explore predictors of vaccine hesitancy, and 3) Describe informational needs of hesitant and non-hesitant workers.

**Methods:** Descriptive statistics were used to present frequencies of vaccine acceptance. Logistic regression was used to explore predictors of vaccine hesitancy based on sociodemographic characteristics, hesitancy to take other vaccines in the past, previous diagnosis of COVID-19, and individual risk perception of getting COVID-19. Chi square analysis was used to compare differences in informational needs between hesitant and non-hesitant individuals across socio-demographic groups.

**Results:** From a sample of 1003 individuals, 439 (44%) reported that they were very likely to get the vaccine, and the rest (66%) reported some level of hesitancy. Older age, Albanian ethnicity, post-secondary school education, previous diagnosis of COVID-19, previous vaccine acceptance of other vaccines, and increased risk perception of COVID-19 infection were all found to be negatively associated with vaccine hesitancy. In particular hesitant individuals, compared to the non-hesitant, wanted to have more information and reassurance that all main international agencies (i.e. FDA, WHO, EMA) were all in accordance in recommending the vaccine and that they would be free to choose if getting the vaccine or not without consequences (p<0.01).

## 1. Introduction

COVID-19 is truly an unprecedented global crisis that has presented unique public health and healthcare challenges to the world. As of December 5, 2021, over 5.2 million people across the globe have died from COVID-19 complications.^1^ In December 2020, biotech companies Pfizer and Moderna released vaccines under emergency use authorization, which governments across the globe prioritized for the elderly, the vulnerable, and essential workers.^2,3^ However, the value of vaccine protection extends only as far as the publics’ willingness to get vaccinated.

Despite the plethora of research proving the effectiveness of vaccines in preventing COVID-19 spread and severe clinical outcomes, vaccine hesitancy remains a challenge in the management of this global health crisis.^4,5,6^ Despite a massive and uninterrupted nationwide vaccination campaign in the Republic of North Macedonia, vaccination coverage remains low compared to western nations. ^7^ As of December 2021, official data from the WHO European COVID-19 vaccination monitor reveal that only 38% of the North Macedonian population is fully vaccinated against COVID-19, and this trend is reflected across other nations in the Balkan peninsula such as Bulgaria (25%), Kosovo (42%), Romania (38%), Serbia (45%), Montenegro (37%), and Croatia (48%)^8^.

The causes contributing to these low vaccination numbers are complex, and studies focused on understanding COVID-19 vaccination acceptance in Eastern European and Balkan countries are limited. Slovenia is the only country for which we could find scientific publications related to COVID-19 vaccine acceptance. Slovenian nursing students showed an acceptance rate of only 33%^8^ and a sample of healthcare workers, not including physicians, showed an acceptance rate lower than the general population (50% versus 57% respectively)^9^. While examining the pre-COVID vaccine literature, a survey of healthcare workers across six European nations showed Bulgarian healthcare workers reporting the greatest vaccine advocacy, uptake, and overall strong positive sentiments for the flu vaccine, compared to other neighboring nations^10^, in contradiction with current COVID-19 vaccination rates of approximately 27%^11^.

The differences in vaccine acceptance patterns across European and Balkan nations are underscored by the sociopolitical intricacies unique to the region. It is presumed that sentiments against vaccinations can be at least partially attributed to an anti-vaccine and anti-government movement that has been growing in this region. These sentiments may have contributed to the European measles outbreak observed in 2018.^12^ Data describing trends in COVID-19 vaccine hesitancy in the Balkans are currently lacking and greatly needed. It is evident that the circumstances preventing vaccination coverage in the Balkan peninsula are unique to each country. Therefore, data that describe the socio-demographic predictors of vaccination sentiments and the informational needs of hesitant individuals are needed to establish effective communication strategies that address vaccine beliefs, attitudes and informational needs that may be related to the spread of misinformation or anti vaccination sentiments on a per country basis.

In this study we focus on the population of the Republic of North Macedonia. To our knowledge this is the first study aimed at understanding COVID-19 vaccine hesitancy in this nation with specific reference to essential workers’ informational needs. This study aims to: 1) Describe rates of vaccine hesitancy, risk perception and sources of COVID-19 information, 2) Explore predictors of vaccine acceptance, and 3) Describe what type of information essential workers would need to make them more likely to accept the vaccine.

## Materials and Methods

### 2.1 Study Design

We conducted a survey of essential workers in North Macedonia to assess their informational needs in regard to the COVID-19 vaccine. A local poll company (Rating Agency) was contracted to conduct phone interviews using the Computer Assisted Telephone Interviewing (CATI) technique. The survey was implemented from May 4-May 16, 2021. A representative sample of the adult population was selected, and screening questions applied to include only respondents that had not taken the vaccine at the time of the survey and were working in job categories prioritized for the vaccination. All data were collected anonymously, and the study conformed to the guidelines and principles of the Declaration of Helsinki. The study protocol and survey instrument were approved by the Harvard T.H. Chan School of Public Health Institutional Review Board.

### 2.2 Survey Instrument

The survey instrument consisted of 35 questions including the following topics/ areas: socio-demographics, acceptance of the COVID-19 and other vaccines, risk perception about contracting COVID-19, experience of COVID-19, health conditions associated with increased risk of severe consequences from the disease, sources of information about the vaccine, and informational needs about the vaccine. Questions were translated from English into Macedonian and back translated into English. A copy of the survey instrument is provided as supplemental material to this manuscript.

### 2.3 Statistical Analysis

We computed descriptive statistics to describe our sample’s socio-demographics and other variables of interest such as: acceptance of the COVID-19 and other vaccines, previous diagnosis of COVID-19 and risk-perceptions, opinions about the government response, and sources of COVID-19 vaccine information. Predictors of vaccine hesitancy were explored by the use of 3 logistic regression models. The dependent variable was derived by the answers to the following question: *“If you were offered a COVID-19 vaccine within two months from now at no cost to you - how likely are you to take it?”*. Answers were coded as 1 (hesitant) = somewhat likely, not interested but would consider it later on, somewhat unlikely, and very unlikely and 0 (not hesitant) = very likely to take it. In model 1, the independent variables include age, sex, and ethnicity. Model 2 includes parameters from model 1 with the addition of education as a proxy for socioeconomic status. Model 3 includes the parameters from model 2 with the addition of previous diagnosis of COVID-19, previous non-COVID-19 vaccine hesitancy, and risk perception of contracting COVID-19.

To describe informational needs about the vaccine we analyzed responses to the following question: *“What would be important for you to know to make you more likely to take the COVID-19 vaccine?”*. Respondents were allowed to select 3 most important topics to them out of 8 possible choices related to vaccine safety and effectiveness, and 3 choices out of 6 related to vaccine policies. Chi Squared test was used test for differences in informational needs between hesitant and non-hesitant individuals. The statistical analysis was conducted by the use of SPSS v.28.

## 3. Results

### 3.1 Sample Characteristics

Table 1 shows the sample characteristics. We gathered responses from 1,003 subjects. Gender is equally distributed with 50% being female. The most represented age group is 35-54 (54%), and 93% have a secondary school education or higher. The most frequently reported income category is 25,000-40,000 denars/month (36%) which is equivalent to $476-$762/month. Most respondents (95%) reported to be employed and 5% of respondents reported being unemployed or volunteers. The most represented job categories are: public health and healthcare workers (23%) grocery store workers (20.4%), food processing workers (12%) and teachers/school staff (11%). In terms of ethnic groups representation, most respondents were Macedonian (77%), followed by Albanian (21%), consistent with the distribution of ethnicity in N. Macedonia. In addition, responses were obtained from all regions: Vardar (68%), Skopje (30%) Polog (15%), Pelagonija (11%) and the Southwest region (11%).

**Table 1.** Sample characteristics and vaccine acceptance (N=1,003)

| Variable |  | n (%) |
| --- | --- | --- |
| Vaccine acceptance |  |  |
|  | I would not take it within 2 months, but maybe later on | 106 (11) |
|  | Very unlikely | 133 (13) |
|  | Somewhat unlikely | 40 (4.0) |
|  | I am not sure | 95 (9.5) |
|  | Somewhat likely | 144 (14) |
|  | Very Likely | 439 (44) |
| Age |  |  |
|  | 18-24 | 56 (5.6) |
|  | 25-34 | 202 (20) |
|  | 35-44 | 275 (27) |
|  | 45-54 | 266 (27) |
|  | 55+ | 204 (20) |
| Gender |  |  |
|  | Male | 496 (50) |
|  | Female | 507 (50) |
| Ethnicity |  |  |
|  | Macedonian | 776 (77) |
|  | Albanian | 207 (21) |
|  | Serb | 7 (0.7) |
|  | Turkish | 5 (0.5) |
|  | Vlach | 3 (0.3) |
|  | Roma | 2 (0.2) |
|  | Bosniak | 2 (0.2) |
|  | Another | 1 (0.1) |
| Education |  |  |
|  | No education | 1 (0.1) |
|  | Primary | 32 (3.2) |
|  | Three-year Secondary | 32 (3.2) |
|  | Secondary | 591 (59) |
|  | Higher Education | 48 (4.8) |
|  | University, Master or PhD | 295 (29) |
| Past Refusal of Non-COVID-19 Vaccines |  |  |
|  | Yes | 117 (12) |
|  | No | 770 (77) |
|  | I don't remember | 78 (7.8) |
|  | I don't Know | 26 (2.6) |
| Previous COVID-19 diagnosis |  |  |
|  | Yes | 247 (25) |
|  | No | 749 (75) |
|  | I don't Know | 7 (0.7) |
| Level of Concern for contracting COVID-19 | At work or outside work | 93 (9.3) |
|  | both at work and outside work | 783 (78) |
|  | no concern | 127 (13) |
| Occupation |  |  |
|  | Unemployed | 6 (0.6) |
|  | Hospital and emergency department workers | 54 (5.4) |
|  | Nursing home, long-term care, and home health care workers | 16 (1.6) |
|  | Public health workers | 79 (7.9) |
|  | Emergency Medical Services workers | 19 (1.9) |
|  | Prisons workers | 4 (0.4) |
|  | Sanitation workers | 15 (1.5) |
|  | Vaccine manufacturing workers | 1 (0.1) |
|  | Other health care workers | 17 (1.7) |
|  | Pharmacy workers | 48 (4.8) |
|  | Teachers and school staff (including childcare and K-12) | 110 (11) |
|  | Food processing workers | 125 (13) |
|  | Grocery store workers | 205 (20) |
|  | Postal and shipping workers | 76 (7.6) |
|  | Public transportation workers | 40 (4) |
|  | Private transportation workers | 70 (7) |
|  | Police or firefighters | 63 (6.3) |
|  | Other first responders | 8 (0.8) |
|  | Volunteer (i.e. CERT, MRC, Red Cross, etc.) | 47 (4.7) |
| Region |  |  |
|  | Vardar | 67 (6.7) |
|  | East | 91 (9.1) |
|  | Southwest | 107 (11) |
|  | Southeast | 87 (8.7) |
|  | Pelagonija | 110 (11) |
|  | Polog | 152 (15) |
|  | Northeast | 85 (8.5) |
|  | Skopje | 304 (30) |
| Average monthly income<br>over last 3 months | Up to 10000 denars | 12 (1.2) |
|  | 10 000 - 18 000 denars | 53 (5.3) |
|  | 18 000 - 25 000 denars | 151 (15) |
|  | 25 000 - 40 000 denars | 362 (36) |
|  | Over 40 000 denars. | 346 (35) |
|  | Refuse to answer | 79 (7.9) |

### 3.2 Acceptance of the COVID-19 and other vaccines

When asked the following question *“If you were offered a COVID-19 vaccine within two months from now at no cost to you - how likely are you to take it?”*: 439 (44%) said they were very likely to take it, 144 (14%) somewhat likely, 106 (11%) were not interested but would consider it later on, 40 (4%) were somewhat unlikely, 133 (13%) were very unlikely and 141 (14%) were not sure or refused to answer. Regarding acceptance of other vaccines, 12% of respondents reported that in the past they had refused a vaccine that was recommended to them by a healthcare worker. The most frequently reported reason for refusing a vaccine in the past was believing it was not necessary (40%). Most respondents (84%) did not get the flu vaccine during the pandemic, when asked why, the most frequently reported reason was that there was no need for it (35%).

### 3.3 Experience of COVID-19 and risk perception

Twenty-five percent of respondents said to have been diagnosed with COVID-19 during the pandemic, 55% of respondents had friends or family members who had tested positive and had no or mild symptoms, 35% did not know of anyone who tested positive, and 25% had friends or family members who experienced severe symptoms. Most respondents were concerned about contracting COVID-19 both at work and outside of work (78%).

### 3.4 Sources of information about the COVID-19 vaccine

Table 2 reflects the sources used to obtain information of the COVID-19 Vaccine. Respondents were asked to select their top sources used, and the three most frequently selected sources were national TV news in Macedonian (72%), social media (54%), and conversations with family, friends and neighbors (47 %). Among respondents that used social media Facebook (79%), YouTube (17%) and Instagram (14%) were the most frequently selected vaccine information sources. When asked if the information received from social media had an impact on their level of confidence about the vaccine, 28% reported that their confidence increased, 10% reported that their confidence decreased, and 27% reported that their confidence did not change. Most of respondents said they did not share information about the vaccine on social media (77%). When surveyed about the trust of information they have gotten so far about the COVID-19 vaccine, 68% reported that they were “somewhat” or “a lot” trustful of the information. Respondents were also asked to select top choices for whom they would trust the most to give them information about the COVID-19 vaccine in the near future. the most frequently selected choices were public health experts (83 %), the respondent’s doctor (54%), and family and friends (25%).

**Table 2.** Sources of information regarding the COVID-19 vaccine

| Variable |  | n (%) |
| --- | --- | --- |
| Top sources of COVID-19 vaccine information |  |  |
|  | National TV news: Macedonian | 723 (72) |
|  | National TV news: Albanian | 196 (20) |
|  | National TV news other language | 20 (2) |
|  | Local TV news: Macedonian | 122 (12) |
|  | Local TV news: Albanian | 47 (4.7) |
|  | Foreign TV News | 158 (16) |
|  | Radio Macedonian | 65 (6.5) |
|  | Radio Albanian | 26 (2.6) |
|  | Radio on another language | 1 (0.1) |
|  | News paper | 35 (3.5) |
|  | Social media | 538 (54) |
|  | Conversation with family/neighbors/friends | 475 (47) |
|  | Employer | 69 (6.9) |
|  | I don't know | 5 (0.5) |
|  | Refuse to answer | 3 (0.3) |
| Received COVID-19 vaccine information from social media |  |  |
|  | Did not | 178 (18) |
|  | I don't Know | 17 (1.7) |
|  | Facebook | 795 (79) |
|  | YouTube | 173 (17) |
|  | Twitter | 20 (2) |
|  | Instagram | 140 (14) |
|  | Tik Tok | 4 (0.4) |
|  | Refuse to answer | 5 (0.5) |
| Confidence in COVID-19 Vaccine was affected by social media information |  |  |
|  | Increased confidence | 286 (29) |
|  | Decreased confidence | 102 (10) |
|  | Did not change my confidence | 275 (27) |
|  | Influenced opinions in other ways | 68 (6.8) |
|  | I am not sure | 93 (9.3) |
|  | Refused to answer | 1 (0.1) |
|  | N/A- did not use | 178 (18) |
| Shared COVID-19 vaccine information on social media |  |  |
|  | Yes | 188 (19) |
|  | No | 775 (77) |
|  | I don't Know | 2 (0.2) |
|  | I don't remember | 38 (3.8) |
| Trust in information received about the COVID-19 vaccine | Not at all | 107 (11) |
|  | Very little | 141 (14) |
|  | Somewhat | 367 (37) |
|  | A lot | 316 (32) |
|  | I don't know | 59 (5.9) |
|  | Refuse to answer | 13 (1.3) |
| Top trusted sources of future COVID-19 vaccine information | Government officials | 172 (17) |
|  | Town leaders (mayor/boards/selectmen) | 36 (3.6) |
|  | Public health experts | 827 (83) |
|  | Employer | 67 (6.7) |
|  | Coworker | 113 (11) |
|  | Primary care doctor | 537 (54) |
|  | Local pharmacy | 93 (9.3) |
|  | Family and Friends | 248 (25) |
|  | Celebrity (sports figure/actor/musician) | 18 (1.8) |
|  | Non-government local leaders (religious leaders/organization) | 22 (2.2) |
|  | I don't know | 21 (2.1) |
|  | Refuse to answer | 3 (0.3) |

### 3.5 Predictors of vaccine hesitancy

Table 3 presents the results of the logistic models. In model 1 female sex, Albanian ethnicity, and older age were found to be inversely associated with vaccine hesitancy. Females have 27% decreased odds of being hesitant compared to male (OR=0.73, 95% CI: 0.56,0.94). Respondents of Albanian ethnicity have 41.8% decreased odds of being hesitant compared to individuals who report Macedonian ethnicity (OR=0.58, 95% CI: .42, .81). Other ethnicities are not significantly predictive of hesitancy. Compared to the youngest age group (18-24) increased age is predictive of decreased vaccine hesitancy, with the oldest age group having 77% decreased odds of being hesitant (OR=0.23, 95% CI: 0.11, 0.46). Model 2 included the parameters from model 1 with the addition of education level, which was used as a proxy for socioeconomic status. When education was included in the regression model, female sex was no longer a significant predictor of vaccine hesitancy. Respondents with more than secondary school education (university, masters programs, and doctorate programs) have 68% decreased odds of being hesitant (OR=0.32, 95% CI: 0.18, 0.58) compared to individuals that had less than a secondary school education. Having a secondary school education is not a significant predictor of hesitancy. Model 3 included parameters from both model 1 and model 2 with the addition of previous non-COVID-19 vaccine hesitancy, previous diagnosis of COVID-19 infection, and degree of concern about contracting COVID-19 (not worried about getting sick, worried about getting sick at home, and/or at work). Respondents reporting that they had avoided a vaccine recommended to them in the past have 48.9% decreased odds of being hesitant to the COVID-19 vaccination compared to individuals who did not avoid them (OR=0.51, 95% CI: 0.33, 0.8).

**Table 3.** Multivariable models of Vaccine Hesitancy Predictors

| Variables | Model 1 |  | Model 2 |  | Model 3 |  |
| --- | --- | --- | --- | --- | --- | --- |
|  | OR | 95% C.I. | OR | 95% C.I. | OR | 95% C.I. |
| 18-24 | *ref* |  |  |  |  |  |
| 25-34 | 0.36 | (0.18, 0.73) | 0.47 | (0.23, 0.95) | 0.46 | (0.22, 0.94) |
| 35-44 | 0.27 | (0.14, 0.54) | 0.34 | (0.17, 0.68) | 0.34 | (0.17, 0.68) |
| 45-54 | 0.28 | (0.14, 0.56) | 0.34 | (0.17, 0.68) | 0.34 | (0.17, 0.70) |
| 55+ | 0.23 | (0.11, 0.46) | 0.28 | (0.14, 0.57) | 0.28 | (0.13, 0.57) |
| Male | *ref* |  |  |  |  |  |
| Female | 0.73 | (0.561, 0.944) | 0.79 | (0.6, 1.03) | 0.85 | (0.65, 1.12) |
| Macedonian | *ref* |  |  |  |  |  |
| Albanian | 0.58 | (0.420, 0.808) | 0.54 | (0.4, 0.79) | 0.5 | (0.35, 0.72) |
| Other | 0.81 | (0.316, 2.078) | 0.77 | (0.3, 1.99) | 0.74 | (0.28, 1.97) |
| Less than secondary school | *ref* |  |  |  |  |  |
| Secondary school |  |  | 0.62 | (0.35, 1.1) | 0.72 | (0.40, 1.3) |
| More than secondary school |  |  | 0.32 | (0.18, 0.58) | 0.4 | (0.22, 0.73) |
| Previous non COVID-19 Vaccine Hesitancy | *ref* |  |  |  |  |  |
| Previous non COVID-19 Vaccine Acceptance |  |  |  |  | 0.51 | (0.36, 0.8) |
| I don't know/ Don't remember/ refuse to answer |  |  |  |  | 0.65 | (0.36, 1.17) |
| Never diagnosed with COVID-19 | *ref* |  |  |  |  |  |
| Diagnosed with COVID-19 in the past |  |  |  |  | 0.68 | (0.5, 0.93) |
| Not concerned with contracting COVID-19 | *ref* |  |  |  |  |  |
| Concerned with contracting COVID-19 at work or outside work |  |  |  |  | 0.41 | (0.21, 0.79) |
| Concerned with contracting COVID-19 at work and outside work |  |  |  |  | 0.24 | (0.14, 0.4) |

Individuals reporting to have been diagnosed with COVID-19 in the past have 32% decreased odds of being hesitant compared to individuals who did not receive a diagnosis (OR=0.68, 95% CI: 0.5, 0.93). Respondents reporting concerns about contracting COVID-19 at either their workplace or home have 59% decreased odds of being hesitant compared to respondents that are not concerned (OR=0.41, 95% CI: 0.21, 0.79). Furthermore, respondents reporting concerns about contracting COVID-19 at both their workplace and their homes have 76% decreased odds of being hesitant to vaccination compared to non-concerned individuals (OR=0.24, 95% CI: 0.14, 0.4).

### 3.7 Informational needs

Respondents were asked what would be important for them to know that would make them more likely to receive the vaccine. Each respondent was asked to choose the three most important topics that were unordered and mutually exclusive. The 3 most frequently selected topics – related to the vaccine safety and effectiveness-for which respondents wanted reassurance were: 1) The vaccine works in stopping the transmission of COVID-19 from one person to another (35%), 2) My risk of getting sick with COVID-19 is bigger than the risk of side effects from the vaccine (33%), and 3) The vaccine works in protecting me from COVID-19 (32%). And 7.2% selected “I don’t know” or refused to answer. The three most frequently selected topics – related to the vaccine policies- for which respondents wanted reassurance were: 1) I will be free to choose if I get the vaccine or not with no consequences (50%), 2) Once vaccinated I will be able to live my life with no restrictions (47%), and 3) Everybody will have equal access to the vaccine regardless of income, race, or insurance status (38%). And 4.6% selected “I don’t know” or refused to answer. Table 4 displays the 3 most frequently selected topics and Figure 1 the differences between hesitant and non-hesitant individuals. In particular hesitant individuals, compared to the non-hesitant, wanted to have more information and reassurance that all main international agencies (i.e. FDA, WHO, EMA) were in accordance in recommending the vaccine and that they would be free to choose if getting the vaccine or not without consequences (p<0.01). The non-hesitant as well expressed the need to have more information about the vaccine. They wanted to know more about the risk benefits of the vaccine – being reassured that getting sick with COVID-19 is bigger than the risk of side effects from the vaccine, that the vaccine will protect them (p<0.01) and that those in charge of approving the vaccine will follow strict rules (p<0.05). They also wanted to be reassured that they would live a life with no restrictions once vaccinated (p<0.01), and that everyone will have equal access to the vaccine (p<0.05).

**Table 4.** Informational needs related to vaccine safety, effectiveness, and policies by vaccine hesitancy status (*= p<0.05, and **= p<0.01) [n=957]

| Informational needs related to vaccine safety and effectiveness |  |
| --- | --- |
| Hesitant individuals (n=518) | 1. The vaccine works in stopping the transmission of COVID-19 from one person to another (35%) *<br>2. The vaccine does not cause immediate or long-term harm (33%)<br>3. The vaccine works in protecting me from COVID-19 (29%)* |
| Non-Hesitant individuals (n=439) | 1. My risk of getting sick with COVID-19 is bigger than the risk of side effects from the vaccine” (40%) **<br>2. The vaccine works in protecting me from COVID-19” (37%)*<br>3. Those approving the vaccines are following strict rules (32%)* |
| Informational needs related to vaccine policies |  |
| Hesitant individuals (n=518) | 1) I will be free to choose if I get the vaccine or not with no consequences (57%) **<br>2) Once vaccinated I will be able to live my life with no restrictions (37%)<br>3) Everybody will have equal access to the vaccine regardless of income, race, or insurance status” (32%). |
| Non-hesitant individuals (n=439) | 1) Once vaccinated I will be able to live my life with no restrictions" (62%)<br>2) Everybody will have equal access to the vaccine regardless of income, race, or insurance status” (44%),<br>3) I will be free to choose if I get the vaccine or not with no consequences (42%). |

**Figure 1.**
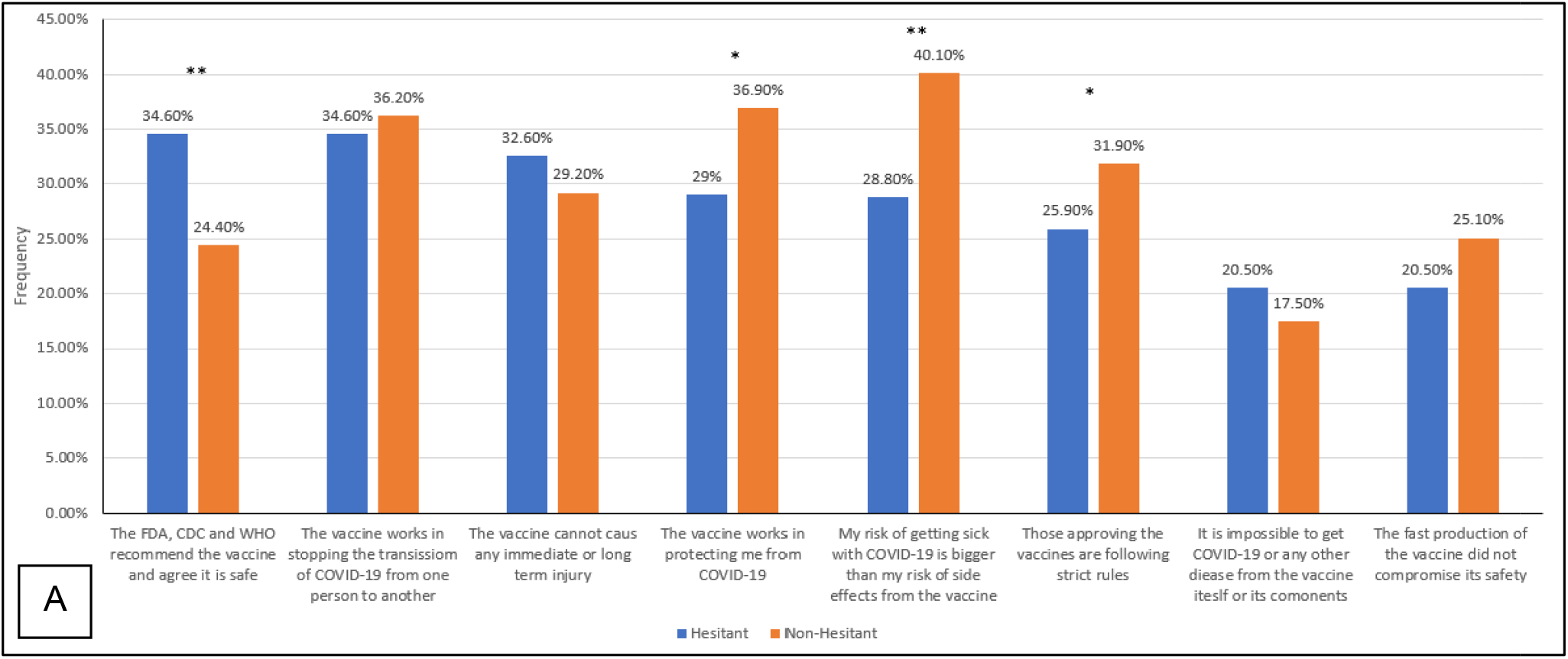
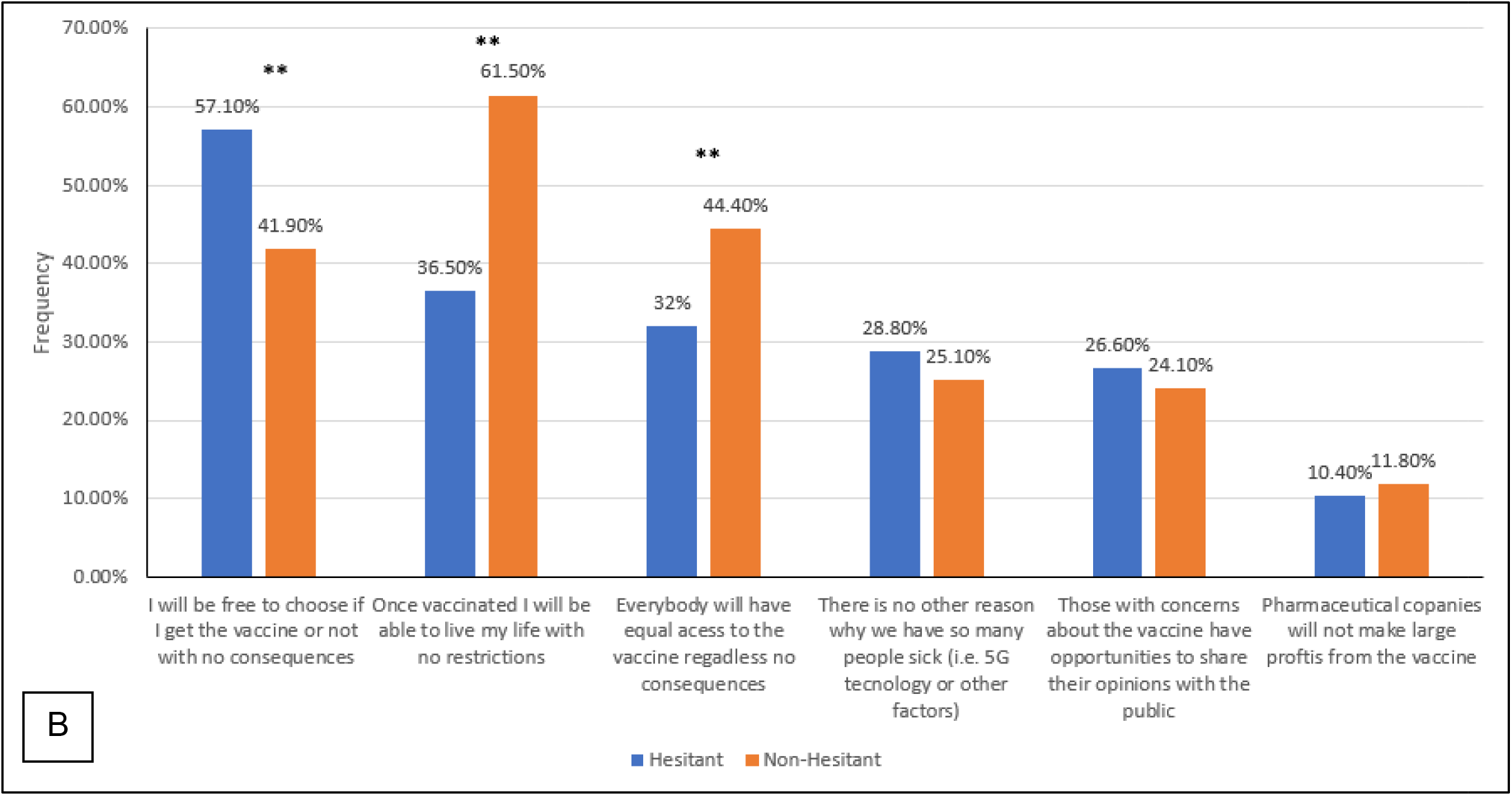
Informational needs of the respondents ranked in descending order for hesitancy: A) Topics related to vaccine safety and efficacy. B) Topics related to vaccination policies (*= p<0.05, and **= p<0.01).

## Discussion

As of December 2021, approximately 40% of adults in the Republic of North Macedonia have been fully vaccinated against COVID-19. The rate is well short of the Western European countries with vaccination rates over 60%. In November 2021, hundreds of citizens protested in Skopje against the Government’s warning for compulsory vaccination for health workers, and the requirement for public sector employees to show a vaccination certificate or negative test on a weekly basis. North Macedonia has struggled with vaccine supply shortages since the early days of the vaccine distribution campaign, the vaccination program began in mid-February and similarly to other countries supplies have been prioritized to healthcare workers, the elderly and essential workers.

Our survey was conducted in May 2021 when approximately 4% of the country 2.1 million residents had been vaccinated. As such the survey was implemented at a time of high demand and low supply, yet our results indicate that vaccine hesitancy may have impacted vaccine uptake. Our survey found approximately 66% of respondents reporting some degree of hesitancy in getting the vaccine, which is consistent with a similar survey conducted in the United States at the start of the vaccine distribution campaign.^13^ When we explored the predictors of such hesitancy previous vaccine avoidance was associated to hesitancy, as well as having been diagnosed with COVID-19 in the past and being concerned of contracting the disease. Both hesitant and non-hesitant individuals reported to need more information about the vaccine, however they had different priorities in terms of what it was important for them to know. The hesitant needed reassurance that the vaccine would stop transmission from person to person and that they would be free to choose if getting the vaccine or not, while those more prone to get vaccinated wanted to have more information about the risk benefits of the vaccine and reassurance that once vaccinated, they could live a life with no restrictions.

As Balkan governments have scrambled to secure coronavirus vaccines, this region has become a breeding ground for anti-vaccine movements. The Balkans has long been a hotbed of misinformation, fueled by low levels of trust in government and other institutions. Regarding potential exposure to misinformation, our survey indicate that about 1 out of 5 respondents believed you can get COVID-19 from the vaccine itself, and that COVID-19 could be caused by 5G. Interestingly, in this case as well these results are not that different from what reported in a similar study conducted in the United States.^13^ Beliefs in unproven theories may be the result of lack of trust in institutions, due to the politicized response to the pandemic. The hampered EU integration has led to skepticism towards Western countries distributing the vaccine, fueling anti-vax movements in a context where the local government has limited capacity to counter misinformation. In this climate, the vaccines and COVID-19 restrictive measures have been spreading as a western-based conspiracy to impose control over the masses.

### Limitations

This study has several limitations. First, our sample is not representative of all essential workers in the Republic of Macedonia. However, a representative sample of essential workers would be very difficult to obtain given the multitude of job categories included in the list of essential workers and the lack of data on how many people work within each category in the nation. Second, the lack of longitudinal data does not allow us to study changes in the willingness to be vaccinated, therefore we do not know if those who were hesitant in May are the same people who are hesitant now, or if the informational needs we identified are reflective of current concerns. Finally, our results might not be comparable to other national polls or surveys because of potential differences in the survey methods, sample populations and questions related to vaccination intent.

## Data Availability

All data produced in the present study are available upon reasonable request to the authors.

