## Supplemental Survey instrument for "Essential Workers’ COVID-19 Vaccine Hesitancy, Misinformation and Informational Needs in the Republic of North Macedonia" (from another version of this preprint: version 1, 2021-12-24)

### VACCINE HESITANCY SURVEY

PI: ELENA SAVOIA


5/5/2021

**Dear Respondent**,

You are invited to take part in a survey conducted by the Emergency Preparedness, Research, Evaluation & Practice (EPREP) program at the Harvard T.H. Chan School of Public Health entitled "**COVID-19 Vaccine Opinions**".

This research is being conducted to learn about people's opinions about the COVID-19 vaccine. **Your participation in this survey is voluntary and you may withdraw your participation at any time for any reason.**

If you take part in this study, you will be asked to talk about your experience during the COVID-19 pandemic and your opinions about the vaccine. The possible risk of participating is answering uncomfortable questions in regards to your experience during the COVID-19 pandemic. You must be 18 or older to participate in this survey.

There are no direct benefits to you in taking part in this research. However, we anticipate that this study will create knowledge that may inform the response to COVID-19.

Your participation in this process will entail completing an online survey (approximately 15 minutes in length). **The survey is anonymous**. None of the information will be reported in any way that will permit you to be identified. If you choose to participate, you may change your mind and leave the study at any time. Refusal to participate or discontinue your participation will involve no penalty or loss of benefits to which you are otherwise entitled. We do not anticipate any reasonably foreseeable risks/discomforts to the prospective participants.

For European participants: In addition to being regulated by applicable U.S. laws, if data about you is collected within the European Economic Area during your participation in this study, that data about you will also be subject to the protections of the General Data Protection Regulation (the "GDPR"). Please note that this survey is anonymous, as such you will not be identified.

This project has been reviewed by the Harvard Longwood Medical Area Institutional Review Board (HMS/HSDM or HCS).

**lick NEXT below** to CONTINUE to complete the survey. Clicking the link below means you give consent to participate. Thank you again for your time and participation.

1. **Have you taken the COVID-19 vaccine?**

○ Yes, I received two or more shots – SCREEN OUT

○ Yes, I received one shot and the vaccine I took requires a second shot

○ Yes, I received one shot and the vaccine I took requires only one shot – SCREEN OUT

○ Not yet, but I have an appointment scheduled for the first dose

○ No, and I do not have an appointment scheduled

2. **What vaccine/s are you eligible to receive? [check all that apply]**

- Pfizer BioNtech
- Moderna
- Johnson & Johson's Jannsen
- Oxford-Astra-Zeneca
- Sputnik V
- Sinopharm (Beijing)
- Covishield (India)
- I do not know

3. **Are you a worker in any of the following categories? [select all that apply]**

❑ Healthcare worker

❑ Residential facility worker

❑ Public health worker

❑ Correctional facilities worker

❑ Vaccine manufacturing or distribution worker

❑ Pharmacy worker

❑ Teacher or school staff

❑ Food processing worker

❑ Grocery store worker

❑ Postal and shipping worker

❑ Transportation worker

❑ Police or firefighter

❑ Volunteer

❑ Other (please specify)

*Tell us about you....*

4. **What is your age category?**

- ○ 18-24
- ○ 25-34
- ○ 35-44
- ○ 45-54
- ○ 55-64
- ○ 65-74
- ○ 75+

5. **Birth Sex?**

- ○ Male
- ○ Female

6. **Please select your region of residence:**
   **Use drop down menu with list**
   - ○ Eastern
   - ○ Northeastern
   - ○ Pelagonia
   - ○ Polog
   - ○ Skopje
   - ○ Southeastern
   - ○ Southwestern
   - ○ Vardar

7. **Are you a citizen?**
    - Yes
    - No

8. **What ethnicity do you consider yourself?**

○ Macedonian

○ Albanian

○ Turk

○ Other (please specify)

9. **What is the highest level of schooling you have completed?**

○ Less than high school

○ High school or equivalent

○ Some college

○ Bachelor's degree

○ Post-graduate degree (i.e. Master, PhD, MD, etc)

10. **In the past 12 months (1 year) are there been occasions in which you were worried about not having enough money or resources to be able to have enough food to eat?**

- Yes
- No

11. **Select the employment status that <u>best describes</u> your current situation [select one option only]:**

○ I am working - paid employee

○ I am working - self-employed

○ I am not working - on unemployment

○ I am not working - on paid leave or furloughed

○ I am not working - searching for work

○ I am retired

○ I am not working - on disability or worker's comp

○ I am not working - and not looking for a job

○ Other (please specify)

12. **Have you received or requested any of the following forms of support since the outbreak of COVID-19**

**pandemic?**

| | Not requested or received | Have requested but have not yet received | Have requested but the request was rejected | Received |
|---|---|---|---|---|
| Unemployment benefit | | | | |
| Wage support (supplement or replacement while still in employment or short-time working schemes) | | | | |
| Paid sick leave or paid care leave (for example, for those who had to self-isolate or take care of children or dependent adults) | | | | |
| State aid to businesses | | | | |
| Other support from public services to help with living expenses or household needs (e.g. benefits, allowances, vouchers, food) | | | | |

13. **Do you have any of the following conditions? [check all that apply]**

- ❑ Cancer
- ❑ Severe allergies
- ❑ Seizures
- ❑ Immunocompromised state due to therapy or disease
- ❑ Autoimmune disease
- ❑ Overweight
- ❑ Obesity
- ❑ Diabetes (type 1 or 2)
- ❑ Cardiovascular disease
- ❑ Hypertension
- ❑ Pulmonary disease
- ❑ Rheumatological condition
- ❑ Pregnancy
- ❑ I do not have any medical condition
- ❑ Other (please specify)

14. * **How concerned are you about any of the following situations?:**

| | Very concerned | Somewhat concerned | Not concerned |
|---|---|---|---|
| **Contracting COVID-19 at work?** (For example: your work settings that is not your home) | ◯ | ◯ | ◯ |
| **Contracting COVID-19 outside of work?** (For example: at the grocery store, when you are using transportation, or in other aspects of your daily life) | ◯ | ◯ | ◯ |
| **Infecting your family or friends with COVID-19?** | ◯ | ◯ | ◯ |

15. **Have you been diagnosed with COVID-19 by a healthcare worker or lab test result?**
    - ❑ Yes, over a year ago
    - ❑ Yes, in the past 12 months
    - ❑ No

16. **Did any of your close family members or friends experience any of the following? [Check all that apply]:**

- ❑ Tested positive for COVID-19 and had no symptoms or mild symptoms
- ❑ Tested positive for COVID-19 and had severe symptoms
- ❑ Died of COVID-19
- ❑ Lost their job or had a salary reduction due to COVID-19
- ❑ Got vaccinated for COVID-19 and My friends and family members who received the vaccine had a positive experience
- ❑ None of my close family or friends experience any of the above situations

17. **What do you think about the number of cases of COVID-19 reported in your country?**

- ○ The number of cases being reported is much lower than the actual number of cases
- ○ The number of cases being reported is much greater than the actual number of cases
- ○ The number of cases being reported is somewhat accurate
- ○ I don't know

**18. Do you feel you are receiving transparent information about the COVID-19 situation from your national government officials?**
    - ❑ Not at all
    - ❑ moderately transparent information
    - ❑ very transparent information
    - ❑ I do not know

19. **In your life, were you ever recommended a vaccine (other than the COVID-19 vaccine) by a healthcare provider that you did not take?**

- ○ Yes
- ○ No
- ○ I do not remember

**20. If you did not take the vaccine (other than COVID-19) that was recommended to you, what was/were the reason(s)? [check all reasons that applied to that situation]**

- ❑ I do not trust vaccines
- ❑ I did not think it was necessary
- ❑ I did not have enough information about the vaccine

- ❏ I did not think the vaccine was effective
- ❏ The vaccine was too expensive
- ❏ It was not logistically convenient to get the vaccine
- ❏ I did not think the vaccine was safe
- ❏ I was concerned about the side effects
- ❏ I had a prior bad experience with vaccinations
- ❏ I was afraid of needles
- ❏ For religious reasons
- ❏ Other reasons (please specify)

21. **Tell us how well the following statements describe your reactions and thoughts:**

| | Very concerned | Somewhat concerned | Not concerned |
|---|---|---|---|
| I feel anxious when I see the number of COVID-19 cases climbing | ◯ | ◯ | ◯ |
| I find the prospect of a vaccine exciting | ◯ | ◯ | ◯ |
| I feel depressed about the uncertainty of how this pandemic will evolve | ◯ | ◯ | ◯ |
| I get upset when I hear contradictory information about COVID-19 | ◯ | ◯ | ◯ |
| I feel stressed when I am unable to plan my life due to COVID-19 | ◯ | ◯ | ◯ |
| I think that taking chances is part of life and so is taking the vaccine | ◯ | ◯ | ◯ |

22. **How much time do you, on average, spend using social media?**

- Everyday for 3 hours or more
- Every day for more than 1 hour but less than 3 hours
- Alternate days
- Not often
- Never

*Information about the vaccine...*

23. **Did  you get information about the COVID-19 vaccine from social media ? [check all that apply]**

- ❑ No, I did not
- ❑ I am not sure
- ❑ Yes - from Facebook
- ❑ Yes - from YouTube
- ❑ Yes - from Instagram
- ❑ Yes - from TikTok
- ❑ Yes - from Twitter
- ❑ Other social media (please specify which one)

*Social media use ...*

**24. [skip logic following q23] Did the information you got from social media change your level of confidence in the COVID-19 vaccine?**

- ○ Increased my confidence in the vaccine
- ○ Decreased my confidence in the vaccine
- ○ Did not change my confidence
- ○ I am not sure
- ○ Did not change my confidence but influenced my opinion in other ways - please specify

**25. Did you ever share information on social media about the COVID-19 vaccine?**

- ○ Yes
- ○ No

26. **When you read or hear something about the COVID-19 vaccine that concerns what source do you rely on for getting additional information? [check all that apply]**

- ❑ Family members
- ❑ My doctor
- ❑ My friends
- ❑ Social media
- ❑ TV
- ❑ I do my own research online
- ❑ I do nothing
- ❑ other ---------------------------------

*Trust in information...*

27. **From what source did you get the most information about the COVID-19 vaccine? Select up to 3 SOURCES:**

1. Local television news (on TV or on the web)
2. Macedonian language national or cable network news (on TV or on the web)
3. Non-Macedonian language television station (on TV or on the web)
4. National newspaper (i.e. New York Times, Wall Street Journal, USA Today on paper or on the web)
5. My town or other local newspaper (on paper or on the web)
6. Non-Macedonian language newspaper (on paper or on the web)
7. Macedonian language radio
8. Non-Macedonian language radio
9. News portal site such as Yahoo! or MSN
10. Website of a government agency
11. Social media
12. Word of mouth
13. Through my employer
14. Other (please specify)

28. **How much do you <u>trust the information</u> you got so far about the COVID-19 vaccine?**

○ Not at all

○ Very little

○ Somewhat

○ A lot

29. **Who would you trust the most to give you information about the COVID-19 vaccine in the near future? Select your <u>TOP 3 choices</u>:**

❏ National officials

❏ Your local government leaders

❏ Public health experts

❏ Your employer

❏ Your co-workers

❏ Your doctor

❏ Your local pharmacy

❏ Your family and friends

❏ Your community health center

❏ A celebrity (for example: a sports figure, actor, or musician)

❏ Local leaders in your community not in government positions (for example: local organizations, religious leaders)

❏ Other (please specify)

*Your opinions ...*

30. **I think that most of the measures taken so far by the Macedonian government to respond to the COVID-19 pandemic have been:**
    - ❑ Just right
    - ❑ Excessive
    - ❑ Not useful
    - ❑ Counter-productive
    - ❑ I don't know

31. **If you were offered a COVID-19 vaccine - at no cost to you- how likely are you to take it?**

- ○ Very likely
- ○ Somewhat likely
- ○ I am not sure
- ○ Somewhat unlikely
- ○ Very unlikely
- ○ I would not take it at the moment but would consider it later on

*Confidence...*

**32. How much do you agree or disagree with the following statements?:**

1 – Strongly disagree

2 – Disagree

3 – Somewhat disagree

4 – Unsure

5 – Somewhat agree

6 – Agree

7 – Strongly agree

- ❑ You cannot get COVID-19 from the vaccine itself
- ❑ There are no toxic ingredients in the vaccine that can harm your health
- ❑ The vaccine cannot mess up your DNA
- ❑ The vaccine cannot cause infertility
- ❑ The vaccine cannot cause other diseases
- ❑ The fast production of the vaccine did not compromise its safety
- ❑ The vaccine is not going to be used by Governments as a tool to limit our civil rights ( right of assembly, right of movement, right of religion, etc.)

33. **How much do you agree or disagree with the following statements?:**

1 – Strongly disagree

2 – Disagree

3 – Somewhat disagree

4 – Unsure

5 – Somewhat agree

6 – Agree

7 – Strongly agree

- ❏ The vaccine will work in protecting me from getting COVID-19
- ❏ By taking the vaccine I will protect my friends and family from getting COVID-19
- ❏ Everyone should get the vaccine to achieve herd immunity
- ❏ Getting COVID-19 is worse than experiencing potential side effects from the vaccine
- ❏ Natural remedies will not protect me from COVID-19

34. **How much do you agree or disagree with the following statements?:**

1 – Strongly disagree

2 – Disagree

3 – Somewhat disagree

4 – Unsure

5 – Somewhat agree

6 – Agree

7 – Strongly agree

- ❏ People should be free to decide if getting vaccinated or not with no consequences for their job or personal life
- ❏ People should have the option to choose the vaccine brand they want
- ❏ People should be allowed to live their life with no restrictions once vaccinated
- ❏ Healthcare professionals and scientists with concerns about the vaccine should have opportunities to share their opinions with the public
- ❏ Everybody should have equal access to the most effective and safe vaccine regardless of income, race, or immigration status
- ❏ There is no elite group that will achieve financial power if people are getting vaccinated
- ❏ There is no microchip with tracking capabilities inserted in the vaccine

35. **If you have other opinions about the vaccine you would like to share please write them here;**

End of survey - **Thank you for your time.**
